# Concurrent cognitive and physical training for cognition in men with prostate cancer: a study protocol for a pilot randomised controlled trial

**DOI:** 10.64898/2025.12.16.25342320

**Authors:** Alanah Pike, Joseph M. Northey, Catherine Paterson, Hollie Speer, Kristy Martin, Nicolas Cherbuin, Amit Lampit, Greg McRoberts, Ganes Pranavan, Ben Rattray

**Affiliations:** UC Research Institute for Sport and Exercise, Building 29, University Drive, University of Canberra, ACT 2617, Australia; Discipline of Sport and Exercise Science, Faculty of Health, University of Canberra, ACT 2617, Australia; Discipline of Nursing, Faculty of Health, University of Canberra, ACT, 2617, Australia; Caring Futures Institute, Flinders University, Adelaide, South Australia, 5042, Australia; Central Adelaide Local Health Network, Adelaide, South Australia, 5000, Australia; Robert Gordon University, Aberdeen, Scotland, United Kingdom; National Centre for Epidemiology and Population Health, Australian National University Canberra, ACT, 2600, Australia; Department of Psychiatry, University of Melbourne, Carlton VIC, 3010, Australia; Prostate Cancer Support Group, Canberra, ACT, 2601, Australia; Medical Oncologist, The Canberra Hospital, Yamba Drive, ACT, 2605 Australia

**Keywords:** androgen deprivation therapy, aerobic exercise, brain games, BrainHQ, cycling, fatigue, intervention, oncology

## Abstract

**Background:** Men with prostate cancer undergoing treatment commonly report poorer cognitive performance than their age-matched controls. Within other populations, modifiable risks factors including cognitive and physical activity can independently provide potent cognitive and neuroprotective benefits. However, greater benefits may be achieved by combining stimuli simultaneously (concurrently) to promote greater neurogenesis, connectivity, and improved cognitive outcomes. This approach has been tested in older populations with and without cognitive impairment, but initial acceptability, feasibility and efficacy are yet to be demonstrated in men affected by prostate cancer. This research aims to 1) demonstrate the acceptability and feasibility of a concurrent cognitive and physical training intervention against pre-set progression criteria and, 2) provide initial estimates of intervention effectiveness on cognitive function, fatigue and quality of life in men with prostate cancer on ADT.

**Methods:** Seventy-six men diagnosed with prostate cancer receiving androgen deprivation therapy will be the recruitment target. This pilot and feasibility study will employ a randomised controlled trial design where participants will be assigned to one of four groups: 1) concurrent training – moderate intensity stationary cycling whilst simultaneously performing computerised cognitive training, 2) moderate intensity stationary cycling only, 3) computerised cognitive training only or 4) wait-list control group. Training will be completed twice a week for 8 weeks, building up to 60 minutes per session. Assessment of the first aim will include an appraisal of recruitment success, adherence, adverse events, subjective acceptability and the principle of no cognitive harm in response to the intervention. In line with aim two, additional measures will include a computerised cognitive battery and subjective questionnaires on cognition, fatigue and quality of life.

**Discussion:** Internationally, many men continue to grapple with the debilitating effects of cognitive decline after a diagnosis and associated treatment with little, or no, evidence-based self-management strategies advised in practice. This timely research will explore the feasibility of concurrent training to treat the cognitive side-effects of androgen deprivation therapy and, in turn, inform the design of a future large-scale clinical trial.

**Trial Registration:** This project was registered with the Australian and New Zealand Clinical Trials Registry: ACTRN12623000767606 on 13^th^ of July 2023.

## Background

Globally, prostate cancer is the second most common cancer among men (1) with rising incidence rates concomitant with an ageing population and improved screening (2, 3). Although the prognosis is favourable in most high-income countries, and the associated mortality rates have continued to fall since the mid 1990’s (4), many men continue to experience severe side-effects from the cancer and associated treatments with little or no self-management advice (5). Specifically, Androgen Deprivation Therapy (ADT) is commonly associated with distressing levels of fatigue (6), coupled with a higher predisposition to cardiovascular disease, diabetes and depression (5, 7). Side-effects such as fatigue have successfully been the target of several intervention strategies including pharmacological aid and varying forms of physical activity (8, 9).

Compared to other side-effects, cognitive disruptions related to prostate cancer and it’s treatment have received less attention in research, but include both objectively assessed deficits and subjective complaints (10, 11). The exact cause of these cognitive disruptions remain unclear, although ADT is often implicated (10). For instance, ADT has been linked to structural (12, 13) and functional changes in the brain (12). In addition, executive functioning has been shown to be impacted by ADT (14, 15) as well as verbal memory (14, 16, 17) attention (14, 16), processing speed (16), visual processing (18) and spatial ability (18). Indeed, there is evidence that the discontinuation of hormone therapy in men with prostate cancer is associated with better cognitive ability for verbal memory tasks (19). Regardless of the specific cause of cognitive side-effects related to prostate cancer treatments, preventing and mitigating these side-effects requires more attention. Given the likely link between structural and functional brain pathways, as well as a higher level of cognition potentially mitigating against excessive cognitive fatigue day to day, interventions that can promote improvement in these areas, like engaging in physical and cognitive activity, are well positioned to support men with prostate cancer in the future.

There is good evidence to support the benefits of physical activity on cognitive function across the lifespan (20, 21), but related research in cancer is largely limited to breast cancer (22, 23). In prostate cancer, the limited evidence suggests that men who report greater levels of physical activity perform better on assessments of memory, attention and executive function (24). Within intervention studies, a scarcity of evidence exists, although 8-weeks of combined aerobic and resistance exercise was found to be associated with improvements in self-reported cognitive function (25). This initial evidence, while limited, aligns with findings in other populations and supports the potential for physical activity to benefit cognitive function in men with prostate cancer. As such, there is a clinical need for physical activity-based intervention studies with objective measures of cognitive function to inform the relative effectiveness of interventions for cognition in this high-risk population.

Another intuitive avenue to target cognitive function is through cognitive training, with recent meta-analytical evidence (26) supporting its use to improve cognitive function in older adults without cancer. An initial pilot study in men with prostate cancer receiving ADT suggests that a computerised cognitive training intervention for 1 hour/day, 5 days/week for 8 weeks can improve reaction time in this population (27). Given the rigorous nature of this intervention, further research is required to determine whether a less intensive and time-consuming intervention that incorporates standard care physical activity recommendations may provide as good or greater benefits to cognition and mitigate cognitive fatigue.

Given the evidence for both cognitive and physical training approaches to benefit cognition, there is understandable clinical interest in whether these interventions can provide additive benefits when delivered in a time efficient, simultaneous (concurrent) manner in this population. Such an approach may not only provide added benefits but ease the time burden on self-management behaviours addressing wellness and quality of life.

Recent meta-analytical evidence highlights that cognitive training, performed concurrently with physical training, can provide superior cognitive benefits for older adults with and without cognitive impairment when compared to other lifestyle interventions alone (28, 29). Concurrent interventions that include cognitive and physical training completed simultaneously are proposed to deliver superior neurogenesis (30) and neuroplasticity (31) in the brain, and improved cognitive outcomes (32) compared to the delivery of these treatments in isolation. These benefits could extend to improvements in cancer-related fatigue, providing low-cost time-efficient opportunities for men with prostate cancer to reduce potential side-effects of the cancer and its treatments. It is unclear however, whether the two interventions can be delivered together and tolerated in this clinical population given the concerns around cancer-related fatigue. Consequently, a key initial step prior to more detailed investigations is to assess the acceptability and feasibility of an intervention that involves concurrent cognitive and physical training (i.e., cycling while completing computerised cognitive tasks) in this population.

Thus, the current study is designed to investigate the acceptability and effectiveness of concurrent cognitive and physical training to alleviate potential cognitive side-effects for men with prostate cancer receiving ADT. The design of this pilot study addresses the following aims 1) investigate the feasibility and acceptability of a concurrent cognitive and physical training intervention in men with prostate cancer against preset progression criteria and 2) provide initial estimates for the effect of a concurrent cognitive and physical training intervention on cognitive function, fatigue and quality of life outcomes in this population. We hypothesise that 1) concurrent cognitive and physical training will be a acceptable and feasible intervention among men diagnosed with prostate cancer receiving ADT and that 2) the concurrent cognitive and physical training intervention will provide positive cognitive, fatigue and quality of life benefits compared to training those modalities in isolation and the wait-list control group.

## Methods

This research will be conducted in the Australia Capital Territory (Australia) at the University of Canberra Cancer Wellness Centre. The intervention will be delivered by a team of Accredited Exercise Physiologists who will supervise all intervention sessions.

### Study Design

The proposed study protocol is an eight-week pilot randomised controlled trial (Figure 1), which will be conducted and reported according to the Consolidated Standards of Reporting Trials (CONSORT) and Standard Protocol Items: Recommendations for Interventional Trials (SPIRIT) guidelines (Figure 2) (33), for pilot and feasibility trials. The CONSORT (Additional File 1) and SPIRIT checklists (Additional File 2) are presented in the supplementary material.

**Figure 1:**
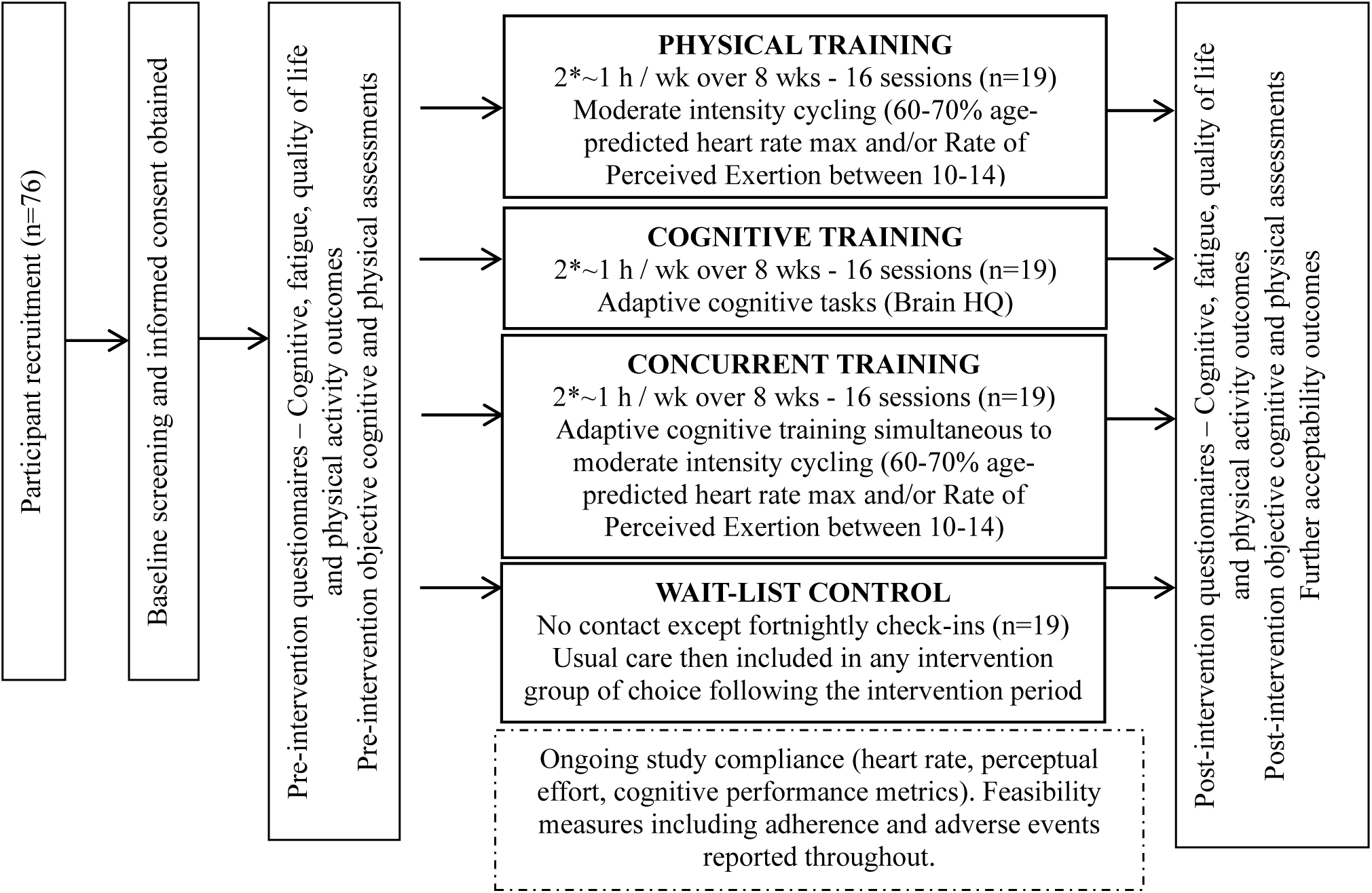
Overview of study design.

**Figure 2:**
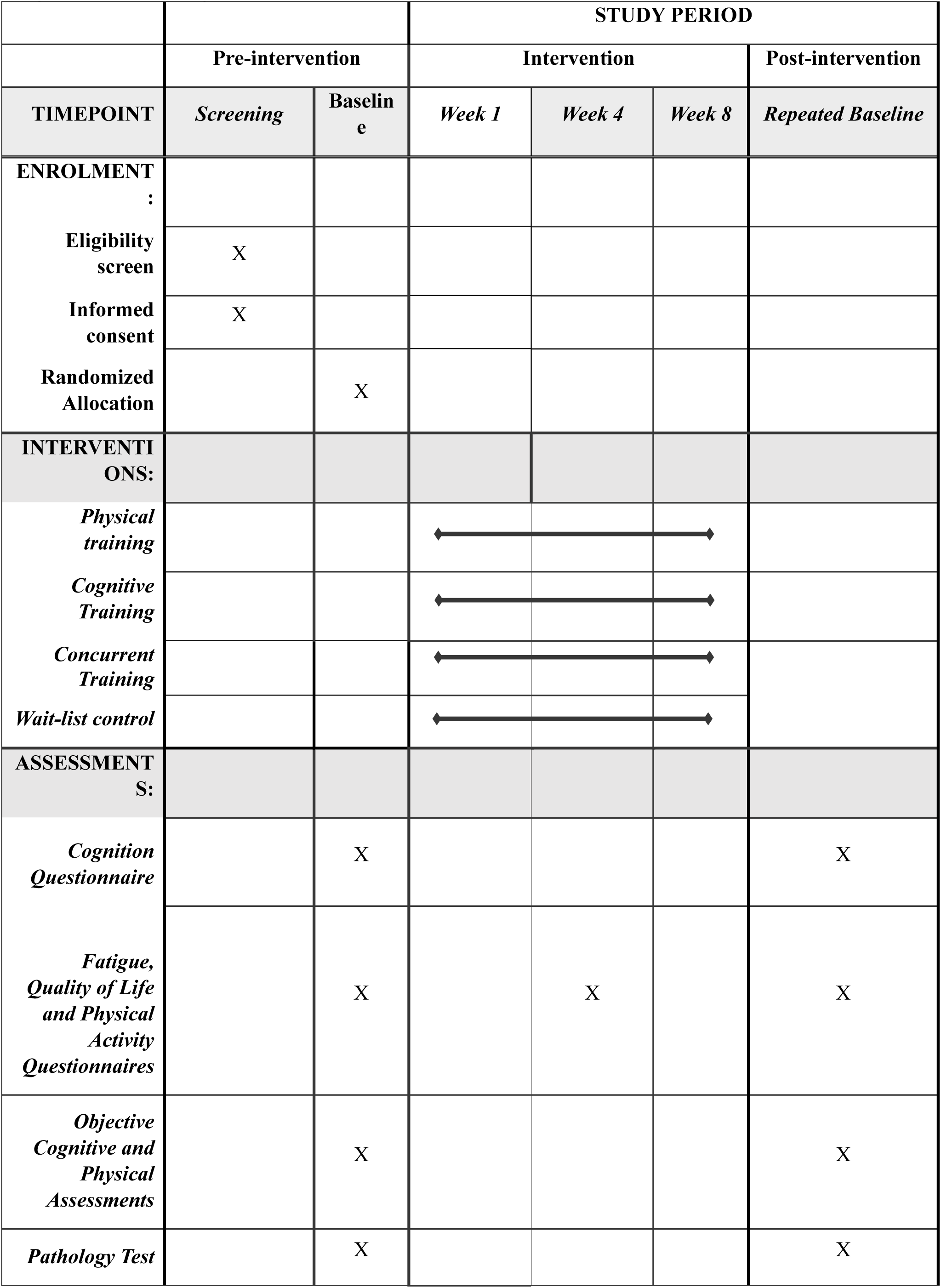
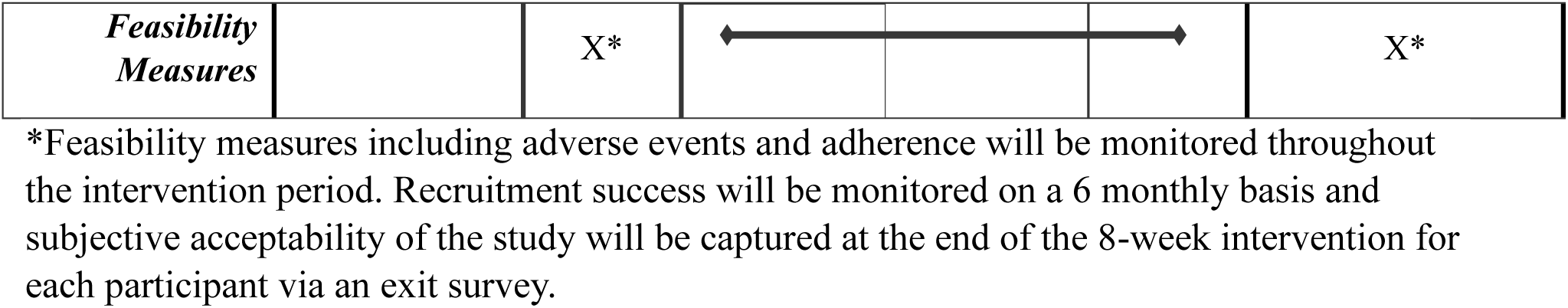
SPIRIT Figure.

The experimental protocol will consist of 1) eligibility screening and informed consent, 2) baseline cognitive health, fatigue, quality of life and physical activity questionnaires, 3) pre-intervention objective cognitive and physical assessments, 4) 8-week training intervention with mid-point questionnaires and 5) post-intervention questionnaires and objective testing, as per Figure 2.

This study has been approved by the University of Canberra Human Research Ethics Committee (#11955). Any amendments to the protocol will be approved by the University of Canberra Human Research Ethics Committee and subsequently submitted to the Australia and New Zealand Clinical Trial registry ACTRN12623000767606 and the funder.

### Participants

The recruitment target is 76 men with prostate cancer over an ∼18-month period. Participants will be recruited from the Australian Capital Territory (ACT) and surrounds utilising the multidisciplinary team’s professional and clinical networks including public and private hospitals. The research team is highly connected to potential recruitment groups. The team includes a Medical Oncologist at The Canberra Hospital and the President of the ACT Prostate Cancer Support Group. The ICON Cancer Centre (co-located with the University of Canberra) will further support recruitment strategies. Outside of these direct routes of participant recruitment, the study will be advertised with flyers in clinics, aged care settings and via media outlets.

The annual incidence of prostate cancer in the ACT is approximately 260 new cases (34). The recruitment target requires a participation rate of 19.5% of the approximately 390 cases expected in the 18-months the study will be completed.

Eligible participants must meet the following inclusion criteria:

- male >18 years old, diagnosed with prostate cancer currently receiving primary ADT for either metastatic or nonmetastatic hormone-sensitive prostate cancer;
- received at least one dose of ADT within the last six months;
- self-assessed reading and writing English proficiency;
- able to provide written informed consent and
- expected life expectancy of more than 12 months.

Based on known interactions with the primary outcome measures, participants will be excluded if 1) they have received chemotherapy or radiation within the last 3 months; 2) they are treated with steroids equivalent to more than 10 mg of prednisolone a day; 3) they have consumed opioid-based analgesics within the last 28 days; 4) they have a history of neurological injury/illness; or 5) they suffer from a concomitant major psychological illness (i.e. bipolar disorder or psychosis).

Participants must be able to complete all aspects of the physical and cognitive intervention. An Adult Pre-Exercise screening will be conducted over the phone in the screening process to confirm eligibility which is verified in an initial baseline familiarisation visit. During this visit, a member of the research team (an Accredited Exercise Physiologist) will confirm eligibility as discussed on the phone in advance, and outline the study procedures, as per the participant information form. The participant is then provided with an opportunity to ask any questions in relation to the study. Participants will be reminded that engagement in the study is completely voluntary, and that they have the right to withdraw at any time, without reason.

If the eligible participant is happy to proceed, they will then be asked to read, and sign a consent form, which will be witnessed and signed by a member of the research team. Following this, the participant will then complete all pre-intervention questionnaires related to demographic health including education level and alcohol intake (Alcohol Use Disorders Identification Tool), as well as subjective cognitive function, fatigue, quality of life (detailed below) and physical activity behaviours (International Physical Activity Questionnaire). Participants are invited to complete these questionnaires on a computer or pen and paper, based on participant preference. Finally, the participant is invited to complete a short familiarisation of the objective computerised cognitive function assessments as an additional screen to confirm sufficient computer literacy. Once complete, participants will attend the laboratory on a separate occasion to complete the full objective cognitive and physical pre-intervention outcome assessments also detailed below.

### Randomisation, group allocation and blinding

All participants who give consent and fulfill the inclusion criteria will be stratified by age (+/-65 years) and then randomly allocated in a 1:1:1:1 ratio to one of four treatment arms: 1) physical training, 2) cognitive training, 3) concurrent cognitive and physical training, or, 4) wait-list control. Randomisation will use a computer-generated sequence of numbers in blocks of variable sizes in a 1:1:1:1 ratio by a researcher not directly involved in the study. After baseline testing (outlined below), a sealed, opaque envelope with the group allocation will be delivered to the participant and training staff. Once opened, the training staff will provide information to the participant about the allocated intervention arm. Blinding of intervention allocation is not possible for any participant or the training staff. Team members completing the post-intervention assessments will be blinded to participant group allocation. Treating clinicians involved in referring potential participants to the study (including author GP) will be blinded to whether potential participants choose to engage with the study.

### Sample size and power calculation

Determining sample sizes in pilot studies typically uses various methods to select samples that are representative of the population of interest. These methods have included less stringent justifications, often applying concepts related to a ‘rule of thumb’. Sample size must ultimately provide sufficient representation of the population to assess feasibility and acceptability. However, it should also provide initial effect estimates to inform progression to larger trials. In line with recommendations for efficient and timely pilot studies (35), we have undertaken a modified power analysis utilising an adjusted alpha (α=0.25).

As a result, we propose to recruit seventy-six participants, which will result in 19 per intervention arm in a 1:1:1:1 ratio. To inform this initial power analysis, we utilised a meta-analysis (28), comparing concurrent training to physical training in older adults. This power analysis was based on a four-arm study, with an effect size of *d*=0.24, 80% power with α=0.25.

### Intervention

The training interventions will consist of 2 sessions per week for eight weeks conducted on non-consecutive days. Irrespective of intervention arm (not including the wait-list control), in Week 1, participants start with 2 x 30-minute sessions, then 2 x 45-minute sessions in Week 2, followed by 2 x 60-minute sessions from Week 3 onwards (inclusive of warm up and cool down), to allow for gradual and more appropriate exposure of the training stimulus.

#### Physical training

The participants will cycle on a recumbent bike ergometer (CXP Target Training Cycle, Matrix Fitness, Australia) for up to 45 minutes at the desired intensity (plus 10-min warm-up and 5-min cool-down) at a fixed intensity. This intensity is relative to their age-predicted heart rate max and rate of perceived exertion (RPE) in accordance with moderate intensity parameters (36) and tolerability as determined by their submaximal exercise assessment. Participants will cycle at a moderate intensity equivalent to 60-70% of their age-predicted heart rate maximum and/or a rate of perceived exertion that corresponds to moderate intensity on the 6-20 Borg scale (11–13) with a one-point buffer on either side (36, 37). The 30-minute sessions in Week 1 consists of a 10-minute warm-up, 15 minutes moderate intensity bout, and 5-minute cool down. Proceeding sessions follow the same warm up and cool-down timeframe, but the bout of moderate intensity exercise in the middle increases accordingly. While cycling, the tablet mounted to the bike displays a YouTube playlist of videos curated with emotionally neutral content at the digression of the participant. This is employed to replicate the attentional demands of the cognitive training without the additional cognitive load.

#### Cognitive training

The participants will complete a series of adaptive multi-domain cognitive tasks (including executive function and working memory tasks) using BrainHQ (Posit Science USA), whilst seated quietly at a desk with minimal distraction. The software adjusts the difficulty to user performance, considering moderators such as learning and fatigue. Each task consists of several levels that broadly increase in complexity in a non-linear manner to maintain a challenging stimulus, by increasing distractors, decreasing target sizes, and reducing response time. Participant performance will be compared against proprietary normative data and required a 70-80% criterion threshold to be met across the training block (typically 10-30 trials) to reveal a new level in the subsequent training session. Lower performance requires participants to repeat the same level on the next attempt. Such multi-domain cognitive training tools provide the most efficacious approach to global and domain specific cognitive benefits (28). The specific cognitive training tools from BrainHQ employed align with domains of cognitive function that may be impacted in this population, which includes: Double Decision (assessing visual processing speed and selective attention), Target Tracker (assessing spatial attention), Hawk Eye (assessing visual speed and attention), Juggle Factor (assessing visuospatial memory and tracking), Scene Crasher (assessing visual working memory) and Eye for Detail (assessing visual processing speed and visual working memory). Participants are given the goal of attaining 20 levels per week of cognitive training from Week 3 onwards (i.e. the maximum exposure of cognitive training).

#### Concurrent cognitive and physical training

The participants will undertake the same cognitive training and physical training interventions described above but delivered simultaneously. *Figure 3* depicts the setup of this intervention. The touchscreen utilised for responses (task-dependent) will be fixed to the stationary bike utilising a customised gooseneck attachment to the handlebar.

**Figure 3:**
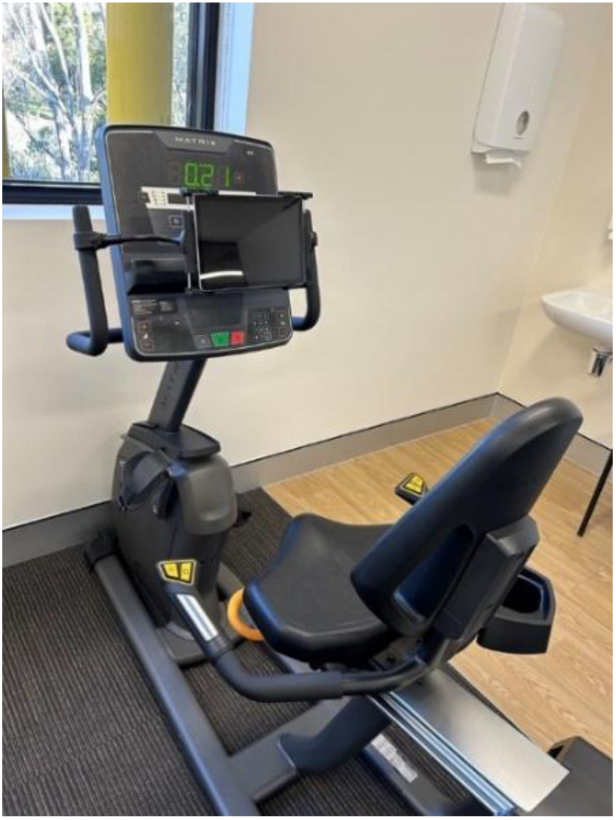
Concurrent training intervention set up showing the attachment of the tablet to the bike.

#### Wait-list control

Participants in the wait-list control will have no contact with the research team outside of a fortnightly check-in over the phone to maintain engagement with the study. The phone calls aim to capture engagement in any kind of physical or cognitive activity within that fortnight, as is described in detail below. Participants will have the option to complete any of the other intervention arms following the initial 8-week intervention period of usual care and subsequent assessments.

### Assessments

The initial session will be used to confirm eligibility, gain consent and complete baseline questionnaires. Following this, participants will attend the laboratory on a separate occasion to complete pre-intervention objective assessments. On arrival, anthropometric measures (height, weight, waist and hip circumference, and blood pressure) will be obtained, followed by the objective cognitive assessments, and the aerobic fitness assessment (both of which are detailed below). A university trained clinical exercise physiologist will monitor all assessments (including electrocardiogram surveillance during the aerobic fitness test), which will act as an additional screening into the trial. Following this assessment procedure, participants will be provided with a referral to attend a local pathology clinic in the ACT to obtain a blood sample as part of the secondary outcome measures. Timing of assessments will consider fatigue, training, and time of day effects within dedicated research spaces at the University of Canberra. Post-intervention assessments will follow the same format after the completion of the 8-week intervention period.

#### Primary Outcomes

Primary outcome measures comprise of feasibility measures in accordance with the first aim. These feasibility measures form the progression criteria, as summarised below.

### Progression Criteria

This pilot clinical trial will provide insights (e.g., feasibility, statistical power, effect estimates) which will guide the development of a larger-scale clinical trial that will aim to improve the cognitive and fatigue outcomes in men affected by prostate cancer receiving ADT. Clinical trial progression criterion as recommended (38), accounts for recruitment success, adherence, adverse events, subjective accounts of the intervention, and the likelihood of benefit (39). *Table 1* provides an overview of the criterion and decision-making related to the progression to a larger trial – each of which are detailed below. As indicated in the progression criteria, further feasibility work may be required depending on the findings of the present study.

**Table 1:** Suggested progression criteria for seeking funding for a larger trial.

|  | **Not to progress to clinical trial** | **Potentially progress with revisions** | **Progress to clinical trial** |
| --- | --- | --- | --- |
| **Recruitment** | Project recruits <50% sample size in designated time frame. Consider alternate designs/qualitative work to understand why. | Project recruits <100% sample size in designated time frame. Evaluate multi-site/locality implications for large trial. Consider qualitative work to understand best recruitment strategies/barriers. | Project recruits designated number in proposed timeframe. Consider locality/ies for larger trial based on power analysis. |
| **Adherence** | Adherence is <50%. | Adherence is 50-65%, consider factors to increase adherence. | Adherence is higher than 65%. |
| **Adverse Events** | Occurrence of any significant adverse events deemed to relate directly to the intervention. | Occurrence minor adverse events (e.g., increased fatigue) consider if protocol changes can be made. | No adverse events reported. |
| **Subjective participant experience** | Subjective accounts are overall negative. | Subjective accounts are neutral. Consider how participant experience could be managed to improve subjective outcomes. | Subjective accounts are overall favourable. |
| **Benefit likelihood** | If results suggest likely harm across outcomes (e.g., cognitive outcomes) > 50% | Results suggest potential harm. Consider changes to protocol. | Results are neutral or positive, proceed to larger trial. |

#### Recruitment success

Recruitment success will be measured by recording recruited participant numbers within the recruitment time frame. Specifics of our recruitment strategies will be described for future reference including details of the most effective recruitment strategies.

#### Adherence

Adherence, retention and withdrawal will be recorded throughout the study. Participants will sign against attendance for training sessions and subsequent session reminders will be provided. Participants will be permitted to make up a “missed” session in the week that the session is missed if there is time to do so (that is, reschedule a session within the same 7-day period).

Sessions throughout the course of the intervention will be booked in advance, and participants will be financially reimbursed in the form of a gift card for their time investment in the study and to assist with retention. The number of withdrawals from the study will be recorded with reasons documented (if provided).

#### Adverse events

When a participant presents for an intervention session, before commencing training, they will be asked about the intervention subjective acceptability (detailed below), as well as whether they have experienced any adverse events in relation to the intervention. Any record of adverse events will be categorised in accordance with the Common Terminology Criteria for Adverse Events (40) as Grade 1 – Mild (asymptomatic or mild symptoms; clinical or diagnostic observation only; intervention not required) through to Grade 5 (death related to adverse event).

Serious adverse events classified as events graded 3, 4 or 5 will be recorded if resulting from the administration of an intervention session within this study and will result in the modification of intervention as appropriate. Events that are serious but are not related to the intervention will not be recorded as serious adverse events.

Alternative causes such as the natural history of the underlying disease, other risk factors and the temporal relationship of the event to the intervention are considered. The Chief Investigator will report serious adverse events related to the intervention (requiring medical or pharmaceutical intervention or hospital admission) to the University of Canberra Human Research Ethics Committee within 24 hours.

#### Subjective Acceptability

Intervention acceptability will be assessed through measures of satisfaction with the intervention, and participants’ attitudes toward the intervention (41). The specific questions included in the exit questionnaire are provided in the additional material (Additional File 3). In summary, participants will be asked for perceptual responses regarding their level of enjoyment of the intervention arm completed and interest and willingness to engage in future interventions. Finally, the survey concludes with an open-ended question allowing participants to comment on their preferred changes to the protocol that would encourage adherence.

Further, prior to participants engaging in any of the intervention sessions involving physical activity, (physical training and concurrent training) the following scales will be used to assess the tolerance of the physical component of the intervention itself: 1) muscle soreness will be captured using a muscle soreness Likert scale from 0 being “A complete absence of soreness” to 6 being “A severe pain that limits my ability to move” and 2) pain will be measured separate to muscles soreness to articulate whether the participant is experiencing pain today, and if so, where. This will be captured using a visual analogue scale ranging from 0 implying no pain and 10 being worst possible pain. We are particularly interested in pain in the lower body given the nature of the intervention arms involved physical training. Participants will be asked to provide a score of the two above scales specifically in response to the previous physical or concurrent training session.

Reporting of physical and cognitive activity outside of the intervention for those within the intervention itself will also be captured pre-, mid- and post- intervention by training staff. Physical activity behaviours will be captured using the International Physical Activity Questionnaire. Cognitive activity is noted from questions including whether they engage in any individual cognitive activities, cognitive activities with others, have occupational cognitive demands or are caring for others. This will help inform the subjective acceptability of the intervention alongside participants’ external life activity.

#### Cognitive function

The benefit likelihood on cognitive function will be assessed pre- and post-intervention in a single sitting, with participants located individually in a quiet room with minimal distractions. Participants will be seated with a laptop and mouse at an individual desk and will be instructed to perform the tasks as quickly and accurately as possible. All tasks were downloaded on 6^th^ April 2022 from the Millisecond (Seattle, USA) test library using the Inquisit Player version 6.5.2 and will be delivered in English (42). Some modifications to the base scripts will be made to be more suitable for this population including changes in timings, trial number and colloquial understandings. Each assessment will include a series of practice trials to be completed before test trials. The cognitive battery takes ∼50 minutes to complete and consists of five tasks in the cognitive domains that have been reported to potentially be impacted by ADT including visuospatial skills, spatial memory, and executive functioning (10). Such domains may also be responsive to concurrent training (28). Category Switch will be the first assessment and will assess executive function through a divided attention task (43). This will be followed by the Visual Simon task assessing executive function, working memory and inhibition (44). The next assessment will be the Groton Maze test assessing immediate and short-term visuospatial memory (45). The second last assessment will be the Visual Search task (Divided Attention) which is a divided attention task assessing visuospatial skills and spatial memory (46). Lastly, the Psychomotor Vigilance test assesses attention, response speed and motor co-ordination (47).

Visuospatial awareness (and fine motor skills) will also be assessed with the Purdue Pegboard. It involves the participant placing as many small pegs as possible into a pegboard in 30 seconds, with both their dominant hand, non-dominant hand, as well as both hands simultaneously. This assessment has specifically been included as visuomotor slowing has been shown in those with advanced prostate cancer (McGinty et al., 2014; Salminen et al., 2004) and may be sensitive to physical activity.

Given the awareness of subjective cognitive complaints, perceived cognitive functioning will be assessed subjectively through the Functional Assessment of Cancer Therapy – Cognitive Function (FACT-Cog) (48). The FACT-Cog is comprised of four subscales: perceived cognitive impairments; perceived cognitive abilities; impact of perceived cognitive impairment on quality of life; and comments from others on cognitive function.

### Secondary Outcomes

Secondary outcomes will include measures of fatigue, quality of life, cardiorespiratory fitness, neurodegeneration blood biomarkers and anthropometric measures assessed pre- and post-intervention.

#### Fatigue

Fatigue is commonly reported in men with prostate cancer and represents a response, and potential side-effect of the intervention. As such, it will be assessed though several means. As an overall intervention outcome, fatigue will be evaluated using the Short-Form 12 (49), including the vitality subscale questions, which will capture subjective general fatigue, pre-intervention, mid-intervention (end of Week 4) and post-intervention.

The NASA Task Load Index will also be employed for participants in all intervention groups for both sessions in Week 1 and Week 8 to provide insight into the perceived mental workload experienced by the participants. The individual dimensions of the NASA Task Load Index will be analysed separately, as research recommends (50). Rate of Perceived Exertion (RPE) will be captured during each of the physical or concurrent training sessions as a measure of perceived physiological fatigue (51).

Further, we anticipate that fatigue in response to the training intervention itself (any arm) will be common, thus the following measure specifically articulates how participants are coping with the intervention. This will be captured before every intervention session (irrespective of intervention arm) using the graded scale according to the Common Terminology Criteria for Adverse Events (40) as categorised below, and reported as a secondary outcome rather than an adverse event:

- Grade 1: Fatigue relieved by rest;
- Grade 2: Fatigue not relieved by rest; limiting instrumental activities of daily living;
- Grade 3: Fatigue not relieved by rest; limiting self-care activities of daily living.

#### Quality of Life

The Expanded Prostate Cancer Index Composite Short Form will be used to capture prostate cancer symptomology, specifically around urinary and bowel function, and sexual health (52). This is a valid and reliable tool for capturing health-related quality of life for those with prostate cancer (53). Further, the Functional Assessment of Cancer Therapy – General (FACT-G) will also be used to measure four domains of health-related quality of life including physical, social, emotional and functional wellbeing (54).

#### Cardiorespiratory fitness

Aerobic fitness will be assessed using the Åstrand Rhyming submaximal exercise test on a recumbent cycle ergometer (Lode, The Netherlands). This protocol is described in detail elsewhere (55), but briefly, this single-stage test commences at 50 watts of resistance, with periodic increases in resistance if heart rate is below 120 beats per minute (56, 57). Throughout the six-minute assessment, the participant is required to maintain a pedal rate of 50 revolutions per minute (56). Heart rate obtained during minute 5 and 6 of the test are averaged together to use in the nomogram and equation to predict VO_2_max (57).

Given the population of interest, and the likelihood of existing factors that influence heart rate including cardiovascular comorbidities and various medications, participants in this study may not be able to achieve the desired heart rate, but their RPE on the 6 to 20 Borg scale should reflect a submaximal intensity between 11-13.

#### Blood Biomarkers

Neurofilament light chains will be measured in plasma samples collected pre- and post-intervention. Neurofilament light chains are a novel biomarker highly specific to neural damage, thought to relate to neurocognitive decline (58). Participants will be able to attend one of several pathology laboratories around the ACT region in the week prior to commencing, and week after completing, the training intervention/wait-list control period.

#### Anthropometric measures

Pre- and post- intervention, bodyweight will be obtained digitally using a Wedderburn TANITA scale to the nearest 0.1kgs, with participants being asked to take of shoes and remove heavy clothing. Height will also be obtained using a SECA model to the nearest centimetre. These measures will be used to calculate body mass index (kilograms of weight/ height in meters squared). Waist/hip circumference will be measured using an anthropometric tape measure in accordance with World Health Organisation guidelines (59). These measures will be used to calculate waist to hip ratio to provide an indication of cardiovascular health in this population. Manual blood pressure will be also obtained using a Littmann Classic stethoscope and Prestige Medical cuff by trained research staff, pre- and post- the 8-week intervention period.

### Statistical Analysis

Quantitative analysis will be conducted in the R statistical package. The mean and standard deviation of the outcome measures pre-, mid- and post-intervention will be calculated for each group. Group differences in baseline characteristics will be assessed with one-way ANOVA or Kruskal-Wallis test (pending normality of data). Data related to the feasibility of the intervention (e.g., adherence and adverse advents) will be reported as frequencies. To estimate intervention effects (i.e., cognitive and fatigue outcomes), Linear Mixed Models with a random intercept fitted for subjects will be implemented using the lme4 package (60), to consider the repeated measures nature of the data and interindividual variability. As per previous work (22), hypothesis testing from pilot studies should be interpreted with caution, as the small sample sizes can result in imprecise estimates and are unlikely to reach statistical significance. Therefore, we will calculate effect sizes for the pairwise changes in primary outcome measures between the experimental conditions. Effect sizes will be calculated as Cohen’s d using the pooled standard deviation of the random effects to account for a Linear Mixed Model structure. Effect sizes will be interpreted as small, moderate, and large based on cut-off points of 0.2, 0.5 and 0.8, respectively.

### Data security and storage

Two separate databases, a research database and a participant database will be utilised in this study to maintain participant confidentiality. The research database will comprise of deidentified demographic information for analysis and all recorded outcome measures (including questionnaires), assessment scores and training data. No other personal information shall be stored in the research database. Each participant will be assigned unique ID code by an individual independent of the research team that is classified based on age stratification 65 years and younger or >66 years. This participant code will be used to collate all data pertaining to that individual. When conducting data collection, this code will be used within the lab, and data will be stored alongside their participant code in the research database.

The participant database will store the participants personal and contact information necessary for safety and contact purposes, as well as the participant code. This will ensure identifiable personal data is not stored with research data. The participant database will be stored in a password protected file and will serve as the key between identifiable information and de-identified code for the study. Only the Chief Investigator (BR) and lead logistics team members will have access to this file.

All members of the listed research team will have access to the research database unless otherwise specified, and data on this server will not be permitted to be shared beyond the research team without appropriate consent. If any additional researchers wish to be granted access to the research database, they will need to be approved on the ethics application prior to access. All team members will be asked to acknowledge in writing before accessing any participant data that they will comply with the National Health and Medical Research Council guidelines in Human Research.

The research database will be stored on secure, password protected University of Canberra (UC OneDrive) servers with access outlined above. All data will be stored for a minimum of 15 years in line with data storage for clinical trials. The use of the research data will be in line with the terms and conditions of the funding body. Any sensitive personal data recorded on physical media will be stored in locked filing cabinets only accessible by authorized personnel.

## Discussion

This protocol describes a novel pilot clinical trial intervention designed to address cognitive function as well as fatigue and quality of life outcomes in men with prostate cancer receiving ADT. Given the novelty of a concurrent cognitive and physical training intervention, this study will investigate the feasibility and assess the acceptability of this approach against predetermined progression criteria. While concurrent cognitive and physical training interventions show promise across several population groups, important questions remain among patients and clinicians around their acceptability within men with prostate cancer to address cancer-related cognitive and fatigue concerns. As such this focal study provides an important step in evaluating the feasibility, and potential of a novel interventional approach to improving cognitive and fatigue outcomes in a population commonly reporting side-effects in this area.

The proposed study aims to address some of the limitations in the existing literature around the interaction between physical activity and cognitive health in this population. Higher levels of self-reported physical activity have been positively associated with cognitive function in men with prostate cancer undergoing ADT (24). This study will not only use self-reported questionnaires of physical activity but implement an objective physical activity intervention to strengthen the evidence around the causal effect of physical activity on cognition in men with prostate cancer. Additionally, an initial feasibility study investigating cognitive training in this population has also shown some promise in select domains of cognition (27), thus, the proposed study will combine cognitive and physical training to potentially maximise cognitive and brain health benefits. Further, several physical activity facilitators specific to this population group will be employed in this study. This includes providing external motivation, opportunities for social interaction and exercise supervised by an Accredited Exercise Physiologist (61).

In terms of study implementation, the stringent exclusion criteria may prove challenging to find participants who have had at least one dose of ADT in the last 6 months and are currently undertaking it in isolation from any other treatment option (i.e. not alongside chemotherapy or radiation). However, this is intentional, as known structural brain changes and cognitive deficits associated with ADT have been shown 6 months post-treatment commencement (13). Furthermore, chemotherapy or radiation treatment separately may adversely affect cognitive function in other cancer populations (62, 63), and this study seeks to evaluate changes outside these known acute impairments on cognition possibly due to these other specific treatment types.

Time constraints may be considered a significant barrier to engaging in physical activity in this population (64). The current study involves participants in the intervention attending the laboratory on 19 separate occasions which may influence session adherence or deter potential participants. Although we will partly address this by reimbursing participants, the aim of the study is to evaluate the effectiveness of a lifestyle intervention that would be more time efficient than its component parts. The tightly controlled prescription and delivery of the intervention is required as a proof-of-concept and is designed to take advantage of current knowledge in exercise and cognitive neuroscience. Potential future translation of concurrent cognitive and physical training would likely include aspects of home-based training using a mix of technologies to aid in broader uptake and adherence.

This research is critical, as despite routine clinical follow-up with healthcare professionals, men affected by prostate cancer receiving ADT continue to grapple for years with the negative effects of cognitive decline and fatigue, with little, or no supported self-management interventions. High prevalence rates, combined with the positive increases in survival rates means attention must turn to understanding and managing the adverse side-effects of prostate cancer and its treatments. As such, the outcomes of this study will inform future clinical trials through cognitive and physical training to address cognitive and fatigue outcomes in men with prostate cancer receiving ADT.

### List of Abbreviations

ACT: Australia Capital Territory
ADT: Androgen Deprivation Therapy
CONSORT: Consolidated Standards of Reporting Trials
FACT-Cog: Functional Assessment of Cancer Therapy – Cognitive
FACT-G: Functional Assessment of Cancer Therapy – General
RPE: Rating of Perceived Exertion
SPIRIT: Standard Protocol Items: Recommendations for Interventional Trials

## Declarations

### Ethics approval and consent to participate

This study has been approved by the University of Canberra Human Research Ethics Committee (#11955). Participation is voluntary and all participants are required to sign a consent form to be involved. It is reiterated to all those who expressed interest that there will be no implications on the type of treatment or care offered to them by the treating clinician (GP) if they chose not to participate in this project.

## Competing interests

The authors declare that they have no competing interests.

## Funding

This project is supported through the international collaborative research partnership between Cancer Australia and World Cancer Research Fund International, with funding provided by Cancer Australia through the PdCCRSi and by Wereld Kanker Onderzoek Fonds through World Cancer Research Fund International. The funder had no contribution towards study design, data collection, management, analysis and interpretation of data, writing the manuscript or the decision to submit this manuscript for publication. This research was also supported by an Australian Government Research Training Program (RTP) Scholarship https://doi.org/10.82133/C42F-K220.

## Authors’ contributions

BR, JN, AL, NC, KM, CP and GP contributed to the conception of the study and collaborated to define the study protocol and criteria. AP and HS will deliver the intervention. All authors contributed to the design of the final study. AP and HS will co-ordinate this trial. AP, and HS will contribute to the acquisition of data. AP will conduct the statistical analysis, and all authors will contribute to the interpretation of data and writing of the final manuscript. AP, BR and JN prepared this protocol paper. All authors provided feedback on drafts of this paper and have read and approved the final manuscript.

## Data Availability

All data to be produced in the present study are available upon reasonable request to the authors

## Acknowledgements

The authors would like to acknowledge Allison Turner for her assistance in recruiting for this project and the ACT Prostate Cancer Support Group for their support in this project.

## Additional Files

Additional File 1 – CONSORT Checklist (.doc) Additional File 2 – SPIRIT Checklist (.doc)

Additional File 3 – Subjective Acceptability Questions (.docx)

## Notes

### Competing Interest Statement

The authors have declared no competing interest.

### Clinical Trial

ACTRN12623000767606

### Funding Statement

This project is supported through the international collaborative research between Cancer Australia and World Cancer Research Fund International, with funding provided by Cancer Australia through PdCCRSi and by Wereld Kanker Onderzoek Fonds (WKOF) as part of the World Cancer Research Fund International grant programme.

This research was also supported by an Australian Government Research Training Program (RTP) Scholarship https://doi.org/10.82133/C42F-K220.

