## Supplemental File 3 for "Concurrent cognitive and physical training for cognition in men with prostate cancer: a study protocol for a pilot randomised controlled trial"

**Additional File 3: Subjective Acceptability Questions in the Exit Questionnaire**

What aspects of the intervention condition you completed did you like/enjoy? Why?

What aspects of the intervention condition you completed did you dislike? Why?

What aspects of the intervention condition you completed did you find most challenging? Why?

Have you noticed any personal benefits from participating in your assigned intervention condition? Please describe the benefits that you have noticed.

Have you experienced any adverse effects from participating I your assigned training intervention? Please describe that adverse effects you have experienced.

Based on your experience, would you recommend the intervention you completed to others? Please explain your answer.

How likely are you to continue with this style of training in the future? Please explain your answer.

Do you have any thoughts/suggestions for how the intervention you completed could be improved? If so, please explain.
